# Associations of habitual glucosamine use with SARS-CoV-2 infection and hospital admission and death with COVID-19: Evidence from a large population based cohort study

**DOI:** 10.1101/2022.09.05.22279621

**Authors:** Meijun Meng, Yanjun Wu, Ruijie Zeng, Dongling Luo, Rui Jiang, Huihuan Wu, Zewei Zhuo, Qi Yang, Jingwei Li, Felix W Leung, Qian Chen, Weihong Sha, Hao Chen

**Affiliations:** Department of Gastroenterology, Guangdong Provincial People’s Hospital, Guangdong Academy of Medical Sciences, Guangzhou 510080, China; The Second School of Clinical Medicine, Southern Medical University, Guangzhou 510515, China; Shantou University Medical College, Shantou 515000, Guangdong, China; Guangdong Cardiovascular Institute, Guangdong Provincial People’s Hospital, Guangdong Academy of Medical Sciences, Guangzhou 510080, China; School of Medicine, South China University of Technology, Guangzhou 510006, China; David Geffen School of Medicine, University of California Los Angeles, Los Angeles 90032, California, USA; Sepulveda Ambulatory Care Center, Veterans Affairs Greater Los Angeles Healthcare System, North Hills 16111, California, USA; Department of Cardiology, Sun Yat-sen Memorial Hospital, Sun Yat-sen University, Guangzhou 510120, China; Guangdong Provincial Key Laboratory of Arrhythmia and Electrophysiology, Guangzhou 510120, China; Guangdong Provincial Key Laboratory of Food, Nutrition and Health, School of Public Health, Sun Yat-sen University, Guangzhou 510080, China

## Abstract

**Objectives:** To assess the association of habitual glucosamine use with coronavirus 2 (SARS-CoV-2) infection, hospital admission, or mortality with Corona Virus Disease-19 (COVID-19) in a large population based cohort.

**Design:** Population based, prospective cohort study.

**Setting:** UK Biobank.

**Participants:** Participants with complete information on habitual glucosamine use and SARS-CoV-2 infection or COVID-19-related outcomes were included. These participants were registered from 2006 to 2010, followed up until 2022 and participated in SARS-CoV-2 tests between 2020 and 2022.

**Main outcome measures:** SARS-CoV-2 infection, COVID-19 hospital admission, and COVID-19 mortality.

**Results:** At baseline, 20,118 (15.9%) of the 126,518 participants reported as habitual glucosamine users. During the median follow-up 12.16 years, there were 53,682 cases of SARS-CoV-2 infection, 2,120 cases of COVID-19 hospital admission and 548 cases of COVID-19 mortality. The multivariate adjusted hazard ratios of habitual glucosamine users to non-users were 1.02 (95% confidence interval [CI] 0.99 to 1.05) for SARS-CoV-2 infection, 0.73 (95% CI 0.63 to 0.85) for COVID-19 hospital admission, and 0.74 (95% CI 0.56 to 0.98) for COVID-19 mortality. The Cox proportional hazard analysis after propensity-score matching yielded consistent results.

**Conclusions:** Habitual glucosamine use seems to be associated with a lower risk of hospital admission and mortality with COVID-19, but not the risk of SARS-CoV-2 infection.

## 1. Introduction

The COVID-19 pandemic caused by the severe acute respiratory syndrome coronavirus 2 (SARS-CoV-2) has contributed to a substantial number of severe cases and mortality worldwide^1^. COVID-19 can trigger cytokine storms in pulmonary tissues, contributing to detrimental pathological features and lethal complications^2 3^. It was estimated that the global in-hospital mortality with coronavirus disease was 15%, and over 18 million people had died due to the COVID-19 pandemic^1 4^.

Glucosamine is an amino monosaccharide that naturally exists in connective tissues, and its oral supplements can be used for the management of osteoarthritis ^5^. It has been estimated that 20% of the adults in the United States use glucosamine ^6^. In the United Kingdom, glucosamine is available as a prescription-only medicine for symptomatic relief. Glucosamine and its related products are demonstrated preclinically to mediate and alleviate the process of inflammation by modulating inflammatory mediators such as nitric oxide and reactive oxygen species ^7-11^. Recent studies have indicated that habitual glucosamine use decreases the risk of cardiovascular events, type 2 diabetes, lung cancer, and all-cause mortality ^12-14^.

To date, the associations between habitual glucosamine use and COVID-19 remain unexplored. Given the anti-inflammatory effect and extensive use of glucosamine and the high prevalence of COVID-19 worldwide, it is critical to evaluate their linkages. Herein, by leveraging the updated and population-based cohort data from the UK Biobank, we aim to evaluate the association between habitual glucosamine use and the risks of SARS-CoV-2 infection and hospital admission and death with COVID-19.

## 2. Methods

### 2.1 Study setting and participants

The UK Biobank was a large prospective study involving 502,413 participants recruited between 2006 and 2010. Participants between the ages of 37 and 73 were invited to one of 22 centers for a baseline assessment and where they completed a detailed touchscreen questionnaires and face-to-face interviews, provided biological samples, and also had a series of physical measurements. Written informed consent was acquired from each participant, and ethical approval was obtained from the North West Multi-Center Research Ethics Committee (approval number: 11/NW/0382, 16/NW/0274, and 21/NW/0157). The current study has been approved under the UK Biobank project 83339.

Data from 502,413 participants were available for our study. We excluded participants with incomplete data on glucosamine-covariate at baseline (n=194,385), incomplete data on the SARS-CoV-2 infection or COVID-19 outcomes (n=152,603) and those with the date of death earlier than January 31, 2020 (n=28,907). In total, our analysis included 126,518 participants (**Figure 1; Supplementary Table S2**).

**Figure 1.**
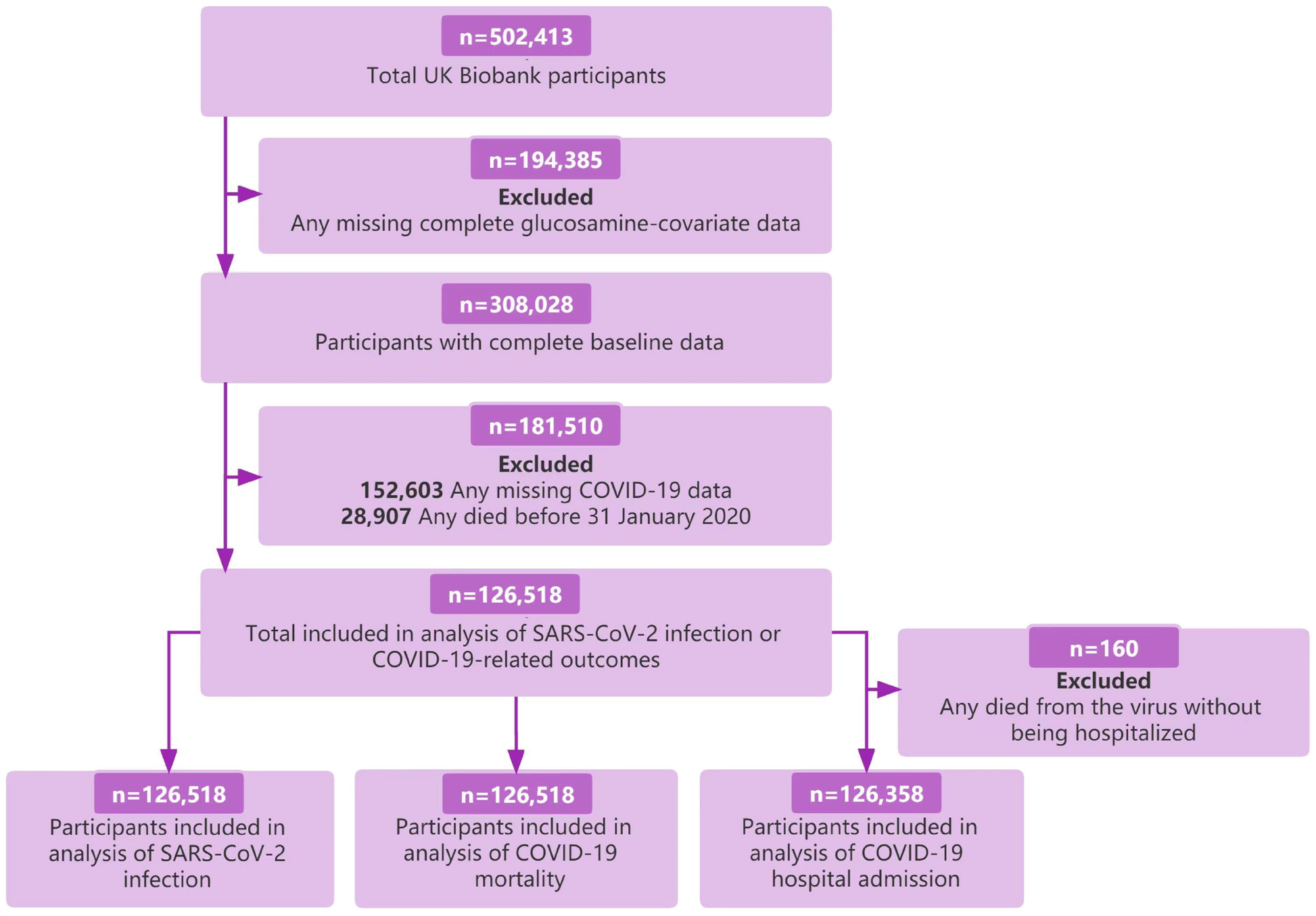
Flow chart of eligible participants selection.

### 2.2 Ascertainment of exposure

The touch screen food frequency questionnaire contained a series of questions: “Do you regularly take any of the following supplements?” Participants could select their answer from a list of supplements, which included glucosamine use (https://biobank.ndph.ox.ac.uk/showcase/field.cgi?id=6179;id=10007;id=20003). From this information, we defined habitual glucosamine usage as 0=no and 1=yes. We only use the assessment of glucosamine supplement use at baseline to perform analyses.

### 2.3 Ascertainment of COVID-19

In this study, the outcomes of SARS-CoV-2 infection (n=53,682), COVID-19 hospital admission (n=2,120) and COVID-19 mortality (n=548) would be focus on. The criteria of SARS-CoV-2 infection was defined as participants with a positive SARS-CoV-2 test and the criteria of COVID-19 hospital admission was defined as ICD-10 code for confirmed COVID-19 (U07.1) or probable COVID-19 (U07.2) in hospital records (excluded those who died from the virus without being hospitalized (n=160)). We defined the fatal outcome as COVID-19 mortality with COVID-19 (ICD-10 U07.1 and U07.2) as the cause of death according to data from the death registers.

### 2.4 Ascertainment of covariates

We used the baseline questionnaire to assess several factors: sociodemographic characteristics (age, sex, ethnicity), socioeconomic status (Townsend Deprivation Index, educational attainment, and household income), lifestyle factors (smoking status, alcohol consumption, body mass index (BMI), dietary intake (vegetables, fruit), comorbidities (hypertension, type 2 diabetes, hypercholesterolemia and arthritis), supplement or co-medication use (antihypertensive drugs, hypolipidemic drugs, insulin, non-aspirin non-steroidal anti-inflammatory drugs [NSAID] and aspirin), vitamin supplementation (vitamin A, vitamin B, vitamin C, vitamin D, vitamin E, multivitamin, or folic acid), and mineral and other dietary supplementation (calcium, iron, zinc, or selenium). These factors were regarded as covariates need to be adjusted in our study, using the directed acyclic graph (DAG) based on existed literature and expert study. (**Supplementary Figure 1**)

The Townsend Deprivation Index, used as an indicator of socioeconomic status, is derived from the residential postcode and is provided directly from the UK Biobank. BMI was calculated as the weight in kilograms (kg) divided by the square of the height in meters (m^2^). Information on comorbidities (hypertension, type 2 diabetes, hypercholesterolemia and arthritis) was collected by self-report at baseline. Prevalent hypertension was defined as a self-reported history of hypertension, the use of antihypertensive drugs, a systolic blood pressure of 140 millimeters of mercury (mmHg) or higher, or a diastolic blood pressure of 90 mmHg or higher. Arthritis was defined by ICD-10 codes (M00-M03, M05-M14).

### 2.5 Statistical analyses

Cox proportional hazards models were used to calculate hazard ratios (HRs) and 95% confidence intervals (CIs) of three different outcomes (SARS-CoV-2 infection, COVID-19 hospital admission, and COVID-19 mortality) according to glucosamine use (yes or no) after confirming whether the assumption of proportional hazards in the Cox regression models was met. It was worth mentioning that the assumption of proportional hazards was fulfilled for main exposure in all outcomes by using Schoenfeld residuals test, which meant the reliability of the Cox proportional hazards model results was high. Three sets of models were used. The non-adjusted model would be set as Model l. Model II was adjusted for age (years) and sex (male or female). The multivariable model (model III) was adjusted for all covariates, including age (years), sex, the Townsend Deprivation Index, ethnicity (white European and others), household income (<£18□000 ($23□500; €21□000), £18□000-£30□999, £31□000-£51□999, £52□000-£100□000, or >£100□000 and “do not know” or missing), educational attainment (college or university degree, non-college or university degree), BMI, fruit consumption (<2.0, 2.0-3.9, or ≥3.9 servings/day), vegetable consumption (<2.0, 2.0-3.9, or ≥3.9 servings/day), smoking status (never, former, or current), alcohol consumption (never, special occasions only, 1-3 times/months, 1-2 times/week, 3-4 times/week, or ≥5 times/week), type II diabetes (yes or no), hypertension (yes or no), hypercholesterolemia (yes or no), arthritis (yes or no), antihypertensive drugs (yes or no), hypolipidemic drugs (yes or no), insulin treatment (yes or no), non-aspirin NSAID drugs use (yes or no), aspirin use (yes or no), vitamin supplementation (yes or no), and mineral and other dietary supplementation (yes or no). Person-years were calculated from the date of baseline recruitment to the date of outcome diagnoses, last follow-up (January 2022 for SARS-CoV-2 infection or COVID-19-related outcomes), whichever came first.

We performed a stratified analysis to assess potential modification effects by the following factors: sex (female or male), age (<60 or ≥60), body mass index (<27 or ≥27), Townsend Deprivation Index (<mean or ≥mean), household income (<52000 or ≥52000),smoking (never, former, or current), alcohol consumption (never, <3 times/week or ≥3 times/week), educational attainment (college or university degree, non-college or university degree), antihypertensive drugs (yes or no), non-aspirin NSAID drugs use(yes or no), type II diabetes (yes or no), hypertension (yes or no), hypercholesterolemia (yes or no), arthritis (yes or no). Simultaneously, the multiplicative interaction between glucosamine use and each covariate on study outcomes were tested, which expressed as *P* for interaction.

Furthermore, we carried out propensity scores matching for glucosamine exposure as an addition to adjust covariates. We matched glucosamine users to non-users at 1:1 ratio using a greedy nearest neighbor method with the MATCHT package in R. The overall quality of the matched sample was assessed by comparing the standardized mean differences (SMD) of all covariates. Among them, the unbalanced covariates between groups (SMD ≥ 0.1) were further adjusted in Cox proportional hazards models to recalculate HRs and 95% CIs. Additionally, the assumption of proportional hazards in the Cox regression models was met here.

We performed all analyses using R software (version 4.2.0, https://www.r-project.org/) in RStudio. We considered a *P* value less than 0.05 (two-sided) to be statistically significant.

### 2.6 Patient and public involvement

No patients were involved in setting the research question or the outcome measures, or in developing plans for design or implementation of the study. No patients were asked to advise on interpretation or writing up of results.

## 3. Results

### 3.1 Population characteristics

**Table 1** shows the baseline characteristics of included participants by glucosamine supplementation usage. The median follow-up was 12.16 (interquartile range [IQR] 11.49-12.82) years. Overall, the mean age of included participants was 55.06 years and 41.2% were male. Among them, 20,118 (15.9%) were glucosamine users, and 106,400 (84%) were glucosamine non-users. Compared with non-users, participants who use glucosamine supplementation were more likely to be female, Caucasians, never smokers, had lower Deprivation index, had medium household income (31,000-51,999£), had three or four times a week alcohol consumption, had college or university degree, had 2-3.9 tablespoons of fresh fruits and raw vegetables per day, and had a lower prevalence of included comorbidities that involved hypertension, type 2 diabetes, hypercholesterolemia and arthritis. Simultaneously, the participants using glucosamine supplementation were also prone to taking vitamin, mineral, and other dietary supplementation, but did not tend to take medicines like antihypertensive drugs, hypolipidemic, insulin, aspirin and non-aspirin NSAIDs.

**Table 1.** Baseline characteristics of UK Biobank participants by habitual glucosamine use.

| Characteristics | Glucosamine<br>non-user | Glucosamine<br>user | Overall |
| --- | --- | --- | --- |
| **Number of participants, n(%)** | 106,400 (84.1) | 20,118 (15.9) | 126,518 (100%) |
| **Age, mean(SD), years** | 54.47 (7.69) | 54.47 (7.69) | 55.06 (7.67) |
| **Sex** |  |  |  |
| Female, n(%) | 61,239 (57.6) | 13,128 (65.3) | 74,367 (58.8) |
| Male, n(%) | 45,161 (42.4) | 6,990 (34.7) | 52,151 (41.2) |
| **Ethnicity** |  |  |  |
| White, n(%) | 102,593 (96.4) | 19,550 (97.2) | 122,143 (96.5) |
| Other, n(%) | 3,807 (3.6) | 568 (2.8) | 4,375 (3.5) |
| **Deprivation index, mean(SD)** | -1.75 (2.82) | -2.06 (2.62) | -1.80 (2.79) |
| **Household income (£)** |  |  |  |
| <18 000, n(%) | 11,345 (10.7) | 2,350 (11.7) | 13,695 (10.8) |
| 18 000-30 999, n(%) | 20,713 (19.5) | 4,736 (23.5) | 25,449 (20.1) |
| 31 000-51 999, n(%) | 28,924 (27.2) | 5,445 (27.1) | 34,369 (27.2) |
| 52 000-100 000, n(%) | 27,386 (25.7) | 4,349 (21.6) | 31,735 (25.1) |
| >100 000, n(%) | 8,077 (7.6) | 1,113 (5.5) | 9,190 (7.3) |
| Missing data | 9,955 (9.4) | 2,125 (10.6) | 12,080 (9.5) |
| **BMI, mean(SD), kg/m<sup>2</sup>** | 26.78 (4.62) | 26.69 (4.42) | 26.77 (4.58) |
| **Alcohol consumption** |  |  |  |
| Daily or almost daily, n(%) | 23,509 (22.1) | 4,931 (24.5) | 28,440 (22.5) |
| Three or four times a week, n(%) | 27,939 (26.3) | 5,585 (27.8) | 33,524 (26.5) |
| Once or twice a week, n(%) | 27,461 (25.8) | 4,900 (24.4) | 32,361 (25.6) |
| One to three times a month, n(%) | 11,745 (11.0) | 2,027 (10.1) | 13,772 (10.9) |
| Special occasions only, n(%) | 9,779 (9.2) | 1,663 (8.3) | 11,442 (9.0) |
| Never, n(%) | 5,967 (5.6) | 1,012 (5.0) | 6,979 (5.5) |
| **Smoking status** |  |  |  |
| Never smoker, n(%) | 62,653 (58.9) | 11,347 (56.4) | 74,000 (58.5) |
| Previous smoker, n(%) | 35,574 (33.4) | 7,740 (38.5) | 43,314 (34.2) |
| Current smoker, n(%) | 8,173 (7.7) | 1,031 (5.1) | 9,204 (7.3) |
| **Education** |  |  |  |
| College or university degree | 58,318 (54.8) | 11,399 (56.7) | 69,717 (55.1) |
| Non-College or university degree | 48,082 (45.2) | 8,719 (43.3) |  |
| **Fresh fruit (tablespoons/day)** |  |  |  |
| <2, n(%) | 34,715 (32.6) | 4,504 (22.4) | 39,219 (31.0) |
| 2-3.9, n(%) | 54,723 (51.4) | 11,326 (56.3) | 66,049 (52.2) |
| ≥3.9, n(%) | 16,962 (15.9) | 4,288 (21.3) | 21,250 (16.8) |
| **Raw vegetable (tablespoons/day)** |  |  |  |
| <2, n(%) | 44,541 (41.9) | 7,197 (35.8) | 51,738 (40.9) |
| 2-3.9, n(%) | 43,782 (41.1) | 8,615 (42.8) | 52,397 (41.4) |
| ≥3.9, n(%) | 18,077 (17.0) | 4,306 (21.4) | 22,383 (17.7) |
| **Supplement or co-medication use:** |  |  |  |
| **Vitamin supplementation, n(%)** |  |  |  |
| Use, n(%) | 28,541 (26.8) | 12,611 (62.7) | 41,152 (32.5) |
| Non-use, n(%) | 77,859 (73.2) | 7,507 (37.3) | 85,366 (67.5) |
| **Mineral and other dietary supplementation, n(%)** |  |  |  |
| Use, n(%) | 31,062 (29.2) | 20,037 (99.6) | 51,099 (40.4) |
| Non-use, n(%) | 75,338 (70.8) | 81 ( 0.4) | 75,419 (59.6) |
| **Antihypertensive drug, n(%)** |  |  |  |
| Use, n(%) | 16,989 (16.0) | 3,274 (16.3) | 20,263 (16.0) |
| Non-use, n(%) | 89,411 (84.0) | 16,844 (83.7) | 106,255 (84.0) |
| **Hypolipidemic drug, n(%)** |  |  |  |
| Use, n(%) | 13,514 (12.7) | 2,651 (13.2) | 16,165 (12.8) |
| Non-use, n(%) | 92,886 (87.3) | 17,467 (86.8) | 110,353 (87.2) |
| **Insulin, n(%)** |  |  |  |
| Use, n(%) | 803 ( 0.8) | 101 ( 0.5) | 904 (0.7) |
| Non-use, n(%) | 105,597 (99.2) | 20,017 (99.5) | 125,614 (99.3) |
| **Aspirin, n(%)** |  |  |  |
| Use, n(%) | 11,110 (10.4) | 2,488 (12.4) | 13,598 (10.7) |
| Non-use, n(%) | 95,290 (89.6) | 17,630 (87.6) | 112,920 (89.3) |
| **Non-aspirin NSAID, n(%)** |  |  |  |
| Use, n(%) | 28,863 (27.1) | 6,559 (32.6) | 35,422 (28.0) |
| Non-use, n(%) | 77,537 (72.9) | 13,559 (67.4) | 91,096(72.0) |
| **Comorbidities:** |  |  |  |
| **Hypertension, n(%)** |  |  |  |
| Yes, n(%) | 23,736 (22.3) | 5,088 (25.3) | 28,824 (22.8) |
| No, n(%) | 82,664 (77.7) | 15,030 (74.7) | 97,694 (77.2) |
| **Type 2 diabetes, n(%)** |  |  |  |
| Yes, n(%) | 5,210 (4.9) | 834 ( 4.1) | 6,044 (4.8) |
| No, n(%) | 101,190 (95.1) | 19,284 (95.9) | 120,474 (95.2) |
| **Hypercholesterolemia, n(%)** |  |  |  |
| Yes, n(%) | 10,337 (9.7) | 2,267 (11.3) | 12,604 (10.0) |
| No, n(%) | 96,063 (90.3) | 17,851 (88.7) | 113,914 (90.0) |
| **Arthritis, n(%)** |  |  |  |
| Yes, n(%) | 7,830 (7.4) | 2,578 (12.8) | 10,408 (8.2) |
| No, n(%) | 98,570 (92.6) | 17,540 (87.2) | 116,110 (91.8) |

BMI: body mass index; NSAID: non-steroidal anti-inflammatory drug; SD: standard deviation

### 3.2 Glucosamine usage and outcomes

The associations for habitual glucosamine use with the risk of SARS-CoV-2 infection, COVID-19 hospital admission, and COVID-19 mortality after Cox proportional hazard analysis were shown in **Table 2**. Within the year recorded by the UK Biobank, we have collected a total of 126,518 participants related to outcome events. Among of them, there were 53,682 participants with all SARS-CoV-2 infection, 2,120 participants with COVID-19 hospital admission, and 548 participants with COVID-19 mortality. In the hazard ratios without adjustment models analysis, we discovered significant inverse associations between glucosamine users and the risk of COVID-19 hospital admission and mortality (all *P*<0.050), but significant positive association between glucosamine users and risk of SARS-CoV-2 infection (*P*<0.050). In addition, in the age and sex-adjusted models, we found inverse correlations that the adjusted hazard ratios associated with glucosamine users were 0.97 (95% CI 0.95 to 1.00; *P*=0.020) for SARS-CoV-2 infection; 0.67 (95% CI 0.58 to 0.76; *P*<0.001) for COVID-19 hospital admission; 0.61(95% CI 0.47 to 0.78; P<0.001) for COVID-19 mortality. Moreover, in the fully adjusted models, including age, sex, ethnicity, deprivation index, household income, BMI, alcohol consumption, smoking status, education, fresh fruit ingestion, raw vegetable ingestion, supplement or co-medication use and history of comorbidities, the adjusted hazard ratios associated with glucosamine users were 0.73 (95% CI 0.63 to 0.85; *P*<0.001) for COVID-19 hospital admission and 0.74 (95% CI 0.56 to 0.98; *P*=0.040) for COVID-19 mortality, which were inverse correlations significantly. However, according to our analysis, there was no significant positive correlation between glucosamine users and risk of SARS-CoV-2 infection (HR 1.02; 95% CI 0.99 to 1.05; *P*=0.120).

**Table 2.** Associations of use of glucosamine with the risk of SARS-CoV-2 infection and COVID-19-related outcomes.

|  | Case/person-years | Non-adjusted model |  | Age and sex-adjusted model |  | Fully adjusted model* |  |
| --- | --- | --- | --- | --- | --- | --- | --- |
|  |  | HR (95% CI) | *P* value | HR (95% CI) | *P* value | HR (95% CI) | *P* value |
| SARS-CoV-2 infection |  |  |  |  |  |  |  |
| Glucosamine non-user | 44,768/543,655 | 1.00 (Reference) |  | 1.00(Reference) |  | 1.00(Reference) |  |
| Glucosamine user | 8,914/108,101 | 1.10(1.07-1.12) | <0.001 | 0.97(0.95-1.00) | 0.020 | 1.02(0.99-1.05) | 0.180 |
| COVID-19 hospital admission |  |  |  |  |  |  |  |
| Glucosamine non-user | 1,856/21,689 | 1.00 (Reference) |  | 1.00(Reference) |  | 1.00(Reference) |  |
| Glucosamine user | 264/3,059 | 0.77(0.67-0.87) | <0.001 | 0.67(0.58-0.76) | <0.001 | 0.73(0.63-0.85) | <0.001 |
| COVID-19 mortality |  |  |  |  |  |  |  |
| Glucosamine non-user | 476/5,586 | 1.00 (Reference) |  | 1.00(Reference) |  | 1.00(Reference) |  |
| Glucosamine user | 72/843 | 0.82(0.64-1.05) | 0.110 | 0.61(0.47-0.78) | <0.001 | 0.74(0.56-0.98) | 0.040 |

CI: confidence interval; HR: hazard ratio
\*Adjusted for age, sex, race (white European, others), Townsend Deprivation Index, average total annual household income (<£18 000, £18 000-£30 999, £31 000-£51 999, £52 000-£100 000, >£100 000, and "do not know" or missing), body mass index, smoking status (never, former, current), alcohol intake, education (with college or university degree, no college degree), fresh fruit consumption (<2.0, 2.0-3.9, or ≥3.9 tablespoons/day), raw vegetable consumption (<2.0, 2.0-3.9, or ≥3.9 tablespoons/day), vitamin supplement use (yes or no), mineral and other dietary supplement use (yes or no), antihypertensive drugs (yes or no), hypercholesterolemia (yes or no), insulin treatment (yes or no), aspirin use (yes or no), non-aspirin NSAID use (yes or no), type 2 diabetes (yes or no), hypertension (yes or no), hypercholesterolemia (yes or no), arthritis (yes or no).

**Supplementary table 1** showed the demographic and clinical characteristics of participants after propensity score-matching (PSM). It listed all the covariates’ standardized mean differences (SMD) were lower than 0.1, which meant well equilibrium and the low difference between groups of our study. Then, the Cox proportional hazard analysis after 1:1 ratio matching indicated consistent results for the outcomes. The hazard ratios associated with glucosamine users were 1.02 (95% CI 0.99 to 1.05; *P*=0.150) for SARS-CoV-2 infection; 0.70 (95% CI 0.60 to 0.82; *P*<0.001) for COVID-19 hospital admission; 0.72 (95% CI 0.53 to 0.97; *P*=0.030) for COVID-19 mortality (**supplementary table 3**).

### 3.3 Interaction and subgroup analyses

We performed subgroup analyses according to potential risk factors (**Figure 2**). A significant multiplicative interaction was observed between glucosamine usage and age on SARS-CoV-2 infection (*P* for interaction=0.001). Meanwhile, the interaction between glucosamine usage and BMI on SARS-CoV-2 infection was also significant (*P* for interaction=0.003). Additionally, we discovered the associations between glucosamine users and the risk of COVID-19 hospital admission outcome had stronger significant multiplicative interactions for alcohol consumption (*P* for interaction=0.010) and educational status (*P* for interaction=0.045). For example, as for COVID-19 hospital admission outcome, the hazard ratios for alcohol consumption at least three times a week was 0.64 (95% CI 0.50 to 0.81), and was 0.68 (95% CI 0.56 to 0.81) for individuals without a college or university degree. Furthermore, the association between glucosamine users and these COVID-19 hospital admission outcome was stronger among alcohol consumption at least three times a week than among alcohol consumption less than three times per week or never. Whereas, the associations between glucosamine users and the risk of COVID-19 hospital admission outcome were not modified by other risk factors, including age, BMI, deprivation index, sex, household income, smoking status, antihypertensive drug use, non-aspirin NSAID use, hypercholestolemia, type 2 diabetes, hypertension, and arthritis. No significant multiplicative interaction was found between glucosamine users and all selected risk factors on fatal COVID-19 outcome (all *P* for interaction >0.050).

**Figure 2.**
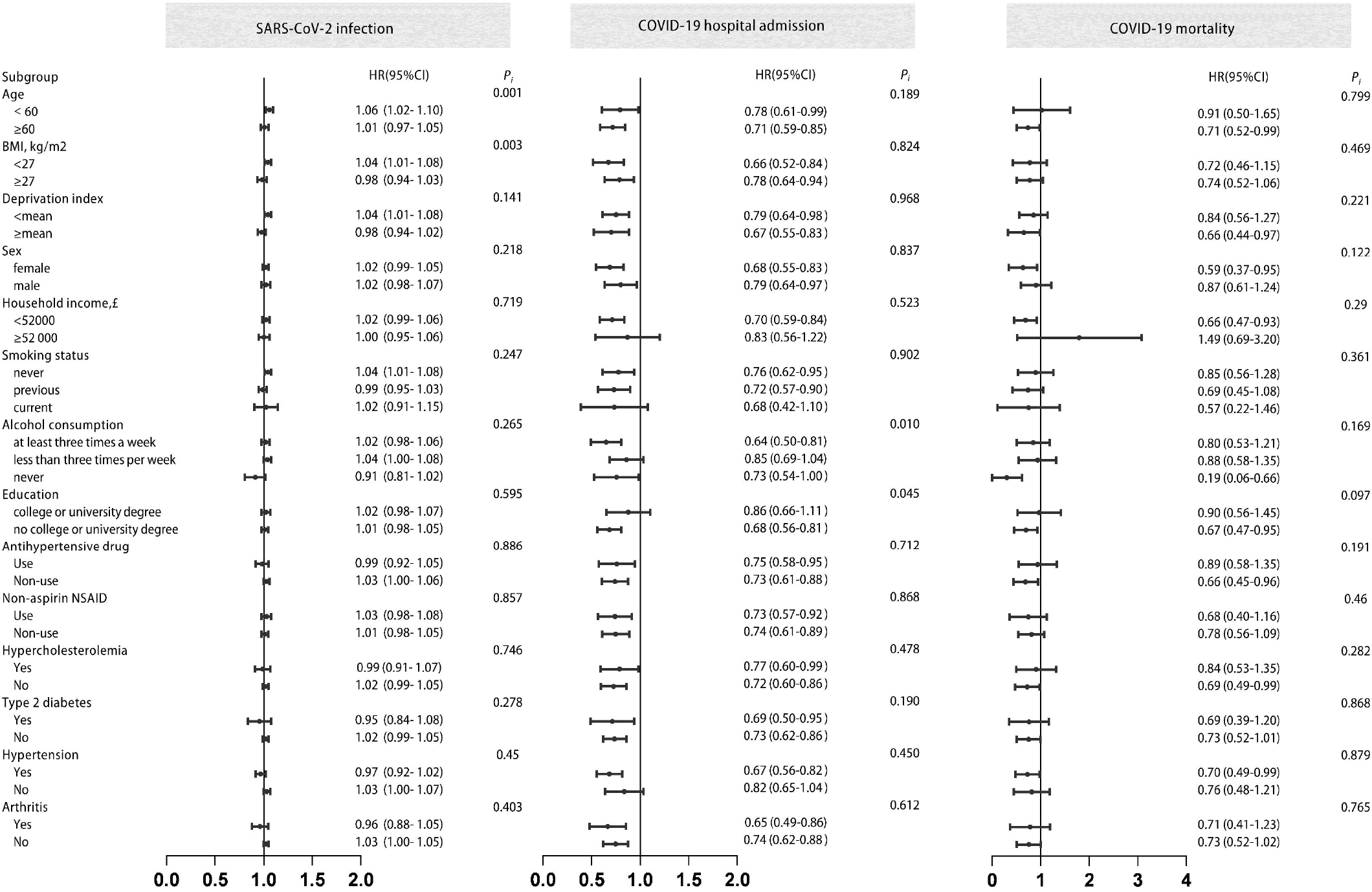
Stratified analysis of glucosamine regular users and risk of SARS-CoV-2 infection and COVID-19-related outcome. Effect estimates were based on age, sex, body mass index (BMI), deprivation index, household income, smoking status, alcohol consumption, education, antihypertensive drug, non-aspirin NSAID, hypercholesterolemia, type 2 diabetes, hypertension, arthritis, using the fully adjusted model. Cl: confidence interval; HR hazard ratio P:P value for interaction.

## 4. Discussion

In this large population-based cohort study, habitual use of glucosamine is associated with a 30% decreased risk of hospital admission and a 28% reduced risk of death among patients with COVID-19, but not the risk of infection. The associations for COVID-19 hospital admission and mortality are independent of age, gender, body mass index, deprivation index, household income, smoking, education, several medications, hypertension, diabetes, hypercholesterolemia, and arthritis.

### 4.1 Comparison with other studies

To our knowledge, none of the previous literature has evaluated the effects of habitual glucosamine on COVID-19. Although habitual glucosamine use has been identified to reduce the risk of lung cancer, type 2 diabetes, cardiovascular diseases, and all-cause mortality by several observational studies^12-17^, its association with COVID-19 remains unknown. In addition, several studies on other dietary supplements, including vitamin C^18^, vitamin D^19^, and folic acid^20^, did not identify any significantly reduced risks for the COVID-19-related outcomes under their use.

As the first attempt on this field, our study might have critical impacts on future research and clinical practice. In spite of the controversial evidence on the treatment of osteoarthritis by glucosamine ^21^, its supplementation could have additional benefits in improving other outcomes of the users. The reduced risks of hospital admission and fatal COVID-19 emphasize the potential benefits of glucosamine.

### 4.2 Biological plausibility

Several proposed mechanisms could account for the observed protective effect of glucosamine and COVID-19-related outcomes. The activation of immune cells, generation of cytokines and chemokines, as well as the positive pro-inflammatory feedback loops are involved in the inflammatory responses to SARS-CoV-2 ^22-24^. In consequence, controlling the inflammatory response is important in improving the hospital admission of COVID-19. Administration of glucosamine in mice decreases the production of inflammatory cytokines and alleviates systemic inflammation^25^. Glucosamine also alleviates oxidative stress and lung inflammation by inhibiting the reactive oxygen species-sensitive inflammatory signaling ^10^. By contrast, the initial infection of SARS-CoV-2 can trigger, but might not directly involve massive inflammatory responses, and therefore glucosamine use does not alter the susceptibility to COVID-19 positivity.

### 4.3 Strengths and limitations of this study

The major strength of our study is the novelty of our findings. In addition, the use of large-scale longitudinal data from the UK Biobank limits the information bias. With adjusting for a variety of covariates, and verification by both traditional logistic regression and propensity-score matching designs, the robustness of our findings is strengthened.

Several limitations also exist in our study. Lack of information referring to the dose and duration of glucosamine use in the UK Biobank limits the assessment of potential causal relationship. Information on glucosamine supplements is based on self-reported data, which could not be verified by other sources based on the design of UK Biobank. Although a wide range of covariates have been adjusted in our study, the results might still suffer from residual confounding, and it is difficult to fully eliminate the effects of healthier lifestyles possessed by glutamine users. Consequently, our findings should be carefully interpreted, while randomized controlled trials could be conducted to further confirm their association.

## 5. Conclusions

Habitual glucosamine use is associated with reduced risks of hospital admission and death with COVID-19. Further well-designed randomized-controlled trials are warranted to further confirm its benefits.

## Data Availability

All data produced are available online at https://www.ukbiobank.ac.uk/uponapplication.

## Acknowledgement

The authors thank the UK Biobank for the access of data, and this research has been conducted under Application Number 83339.

## Author Contributions

MJM, YJW, RJZ, and DLL contributed equally to this work. HC, WHS, QC, and FWL are senior and corresponding authors who also contributed equally to this study. MJM, YJW, and RJZ contributed to data extraction, data analyses, and manuscript drafting. DLL contributed to data interpretation and manuscript drafting. RJ, HHW, ZWZ, QY, and JWL contributed to manuscript drafting. HC, WHS, QC, and FWL contributed to study design, data interpretation, and final approval of the manuscript. The corresponding author attests that all listed authors meet authorship criteria and that no others meeting the criteria have been omitted.

## Figure Legends

**Supplementary Figure 1. Directed acyclic graph (DAG) for evaluation of covariates in the Cox proportional hazard analysis model**. Nodes and arrows respectively represent variables and causal associations. The exposure (Glucosamine) is denoted by a red node, and the outcome (SARS-CoV-2 infection or COVID-19-related outcomes) is labeled by a blue node. The proposed causal associations were based on existed literature and expert study. BMI: body mass index; NSAID: Non-steroidal Anti-inflammatory Drugs.

