## Supplementary Files for "Associations of habitual glucosamine use with SARS-CoV-2 infection and hospital admission and death with COVID-19: Evidence from a large population based cohort study"

**Table S1. The demographic and clinical characteristics of participants after propensity score-matching**

| Characteristics | Glucosamine non-user | Glucosamine user | Overall | SMD |
|---|---|---|---|---|
| **Number of participants, n(%)** | 20,118(50.0) | 20,118(50.0) | 40,236 | |
| **Age, mean(SD), years** | 58.08 (6.93) | 58.19 (6.74) | 58.14 (6.84) | 0.016 |
| **Sex** | | | | 0.013 |
| Female, n(%) | 13,001 (64.9) | 13,128 (65.3) | 26,129 (64.9) | |
| Male, n(%) | 7,117 (35.4) | 6,990 (34.7) | 14,107 (35.1) | |
| **Ethnicity** | | | | 0.009 |
| White, n(%) | 19,519 (97.0) | 19,550 (97.2) | 39,069 (97.1) | |
| Other, n(%) | 599 (3.0) | 568 (2.8) | 1,167 (2.9) | |
| **Deprivation index, mean(SD)** | -2.06 (2.65) | -2.06 (2.62) | -2.05 (2.63) | 0.012 |
| **Household income (£)** | | | | 0.016 |
| <18 000, n(%) | 2,412 (12.0) | 2,350 (11.7) | 4,762 (11.8) | |
| 18 000-30 999, n(%) | 4,649 (23.1) | 4,736 (23.5) | 9,385 (23.3) | |
| 31 000-51 999, n(%) | 5,388 (26.8) | 5,445 (27.1) | 10,833 (26.9) | |
| 52 000-100 000, n(%) | 4,374 (21.7) | 4,349 (21.6) | 8,723 (21.7) | |
| >100 000, n(%) | 1,144 ( 5.7) | 1,113 ( 5.5) | 2,257 ( 5.6) | |
| Missing data | 2,151 (10.7) | 2,125 (10.6) | 4,276 (10.6) | |
| **BMI, mean(SD), kg/m2** | 26.63 (4.52) | 26.69 (4.42) | 26.66 (4.47) | 0.015 |
| **Alcohol consumption** | | | | 0.008 |
| Daily or almost daily, n(%) | 4,886 (24.3) | 4,931 (24.5) | 9,817 (24.4) | |
| Three or four times a week, n(%) | 5,610 (27.9) | 5,585 (27.8) | 11,195 (27.8) | |
| Once or twice a week, n(%) | 4,898 (24.3) | 4,900 (24.4) | 9,798 (24.4) | |
| One to three times a month, n(%) | 2,051 (10.2) | 2,027 (10.1) | 4,078 (10.1) | |
| Special occasions only, n(%) | 1,644 ( 8.2) | 1,663 ( 8.3) | 3,307 ( 8.2) | |
| Never, n(%) | 1,029 ( 5.1) | 1,012 ( 5.0) | 2,041 ( 5.1) | |
| **Smoking status** | | | | 0.006 |
| Never smoker, n(%) | 11,389 (56.6) | 11,347 (56.4) | 22,736 (56.5) | |
| Previous smoker, n(%) | 7,683 (38.2) | 7,740 (38.5) | 15,423 (38.3) | |
| Current smoker, n(%) | 1,046 ( 5.2) | 1,031 ( 5.1) | 2,077 ( 5.2) | |
| **Education** | | | | 0.009 |
| College or university degree | 11,490 (57.1) | 11,399 (56.7) | 22,889 (56.8) | |
| No college or university degree | 8,628 (42.9) | 8,719 (43.3) | 17,347 (43.2) | |
| **Fresh fruit (tablespoons/day)** | | | | 0.007 |
| <2, n(%) | 4,558 (22.7) | 4,504 (22.4) | 9,062 (22.5) | |
| 2-3.9, n(%) | 11,316 (56.2) | 11,326 (56.3) | 22,642 (56.3) | |
| ≥3.9, n(%) | 4,244 (21.1) | 4,288 (21.3) | 8,532 (21.2) | |
| **Raw vegetable (tablespoons/day)** | | | | 0.008 |
| <2, n(%) | 7,276 (36.2) | 7,197 (35.8) | 14,473 (36.0) | |
| 2-3.9, n(%) | 8,568 (42.6) | 8,615 (42.8) | 17,183 (42.7) | |
| ≥3.9, n(%) | 4,274 (21.2) | 4,306 (21.4) | 8,580 (21.3) | |
| **Supplement or co-medication use：** | | | | |
| **Vitamin supplementation, n(%)** | | | | 0.015 |
| Use, n(%) | 12,462 (61.9) | 12,611 (62.7) | 25,073 (62.3) | |
| Non-use, n(%) | 7,656 (38.1) | 7,507 (37.3) | 15,163 (37.7) | |
| **Mineral and other dietary supplementation, n(%)** | | | | <0.001 |
| Use, n(%) | 20,037 (99.6) | 20,037 (99.6) | 40,074 (99.6) | |
| Non-use, n(%) | 81 ( 0.4) | 81 ( 0.4) | 162 (0.4) | |
| **Antihypertensive drug, n(%)** | | | | 0.005 |
| Use, n(%) | 3,312 (16.5) | 3,274 (16.3) | 6,586 (16.4) | |
| Non-use, n(%) | 16,806 (83.5) | 16,844 (83.7) | 33,650 (83.6) | |
| **Hypolipidemic drug, n(%)** | | | | 0.005 |
| Use, n(%) | 2,686 (13.4) | 2,651 (13.2) | 5,337 (13.3) | |
| Non-use, n(%) | 17,432 (86.6) | 17,467 (86.8) | 34,899 (86.7) | |

| | | | | |
|---|---|---|---|---|
| **Insulin, n(%)** | | | | 0.001 |
| Use, n(%) | 99 (0.5) | 101 (0.5) | 200 (0.5) | |
| Non-use, n(%) | 20,019 (99.5) | 20,017 (99.5) | 40.036 (99.5) | |
| **Aspirin, n(%)** | | | | 0.004 |
| Use, n(%) | 2,515 (12.5) | 2,488 (12.4) | 5,016 (12.5) | |
| Non-use, n(%) | 17,603 (87.5) | 17,630 (87.6) | 35,220 (87.5) | |
| **Non-aspirin NSAID, n(%)** | | | | 0.024 |
| Use, n(%) | 6,335 (31.2) | 6,559 (32.6) | 12,894 (32.0) | |
| Non-use, n(%) | 13,783 (68.5) | 13,559 (67.4) | 27,342 (68.0) | |
| **Comorbidities：** | | | | |
| **Hypertension, n(%)** | | | | <0.001 |
| Yes, n(%) | 5,084 (25.3) | 5,088 (25.3) | 10,172 (25.3) | |
| No, n(%) | 15,034 (74.7) | 15,030 (74.7) | 30,064 (74.7) | |
| **Type 2 diabetes, n(%)** | | | | 0.007 |
| Yes, n(%) | 861 (4.3) | 834 (4.1) | 1,695 (4.2) | |
| No, n(%) | 19,275 (95.7) | 19,284 (95.9) | 38,541 (95.8) | |
| **Hypercholesterolemia, n(%)** | | | | 0.001 |
| Yes, n(%) | 2,263 (11.2) | 2,267 (11.3) | 4,530 (11.3) | |
| No, n(%) | 17,855 (88.8) | 17,851 (88.7) | 35,706 (88.7) | |
| **Arthritis, n(%)** | | | | 0.035 |
| Yes, n(%) | 1,922 (9.6) | 2,135 (10.6) | 4,057 (10.1) | |
| No, n(%) | 18,196 (90.4) | 17,983 (89.4) | 36,179 (89.9) | |

BMI: body mass index; NSAID: non-steroidal anti-inflammatory drug; SD: standard deviation; SMD: standardized mean difference.

**Table S2. The numbers (percentages) of participants with missing covariates**

| Covariates | n | % |
|---|---|---|
| Age | 44,690 | 14.51% |
| Ethnicity | 33,289 | 10.81% |
| Deprivation index | 626 | 0.20% |
| Household income | 6,014 | 1.95% |
| Smoking status | 45,873 | 14.89% |
| Alcohol consumption | 1,502 | 0.49% |
| BMI | 38,557 | 12.52% |
| Education | 95,391 | 30.97% |
| Fresh fruit | 19,117 | 6.21% |
| Raw vegetable | 31,385 | 10.19% |
| Vitamin supplementation | 7,337 | 2.38% |
| Mineral and other dietary supplementation | 35,192 | 11.42% |
| Antihypertensive drug | 3,887 | 1.26% |
| Aspirin | 5,963 | 1.94% |
| Non-aspirin NSAID | 5,963 | 1.94% |

BMI: body mass index; NSAID: non-steroidal anti-inflammatory drug

**Table S3. Propensity score-matched analysis**

| | Case/person-years | HR (95%CI) | *P* value |
|---|---|---|---|
| **SARS-CoV-2 infection** | | | |
| **Glucosamine non-user** | **9,077/110,158** | **1.00 (Reference)** | |
| **Glucosamine user** | **8,914/108,101** | **1.02(0.99-1.05)** | **0.150** |
| **COVID-19 hospital admission** | | | |
| **Glucosamine non-user** | **381/4,420** | **1.00 (Reference)** | |
| **Glucosamine user** | **264/3,060** | **0.70(0.60-0.82)** | **<0.001** |
| **COVID-19 mortality** | | | |
| **Glucosamine non-user** | **103/1,199** | **1.00 (Reference)** | |
| **Glucosamine user** | **72/843** | **0.72(0.53-0.97)** | **0.030** |

CI: confidence interval; HR: hazard ratio.

Supplementary Figure 1

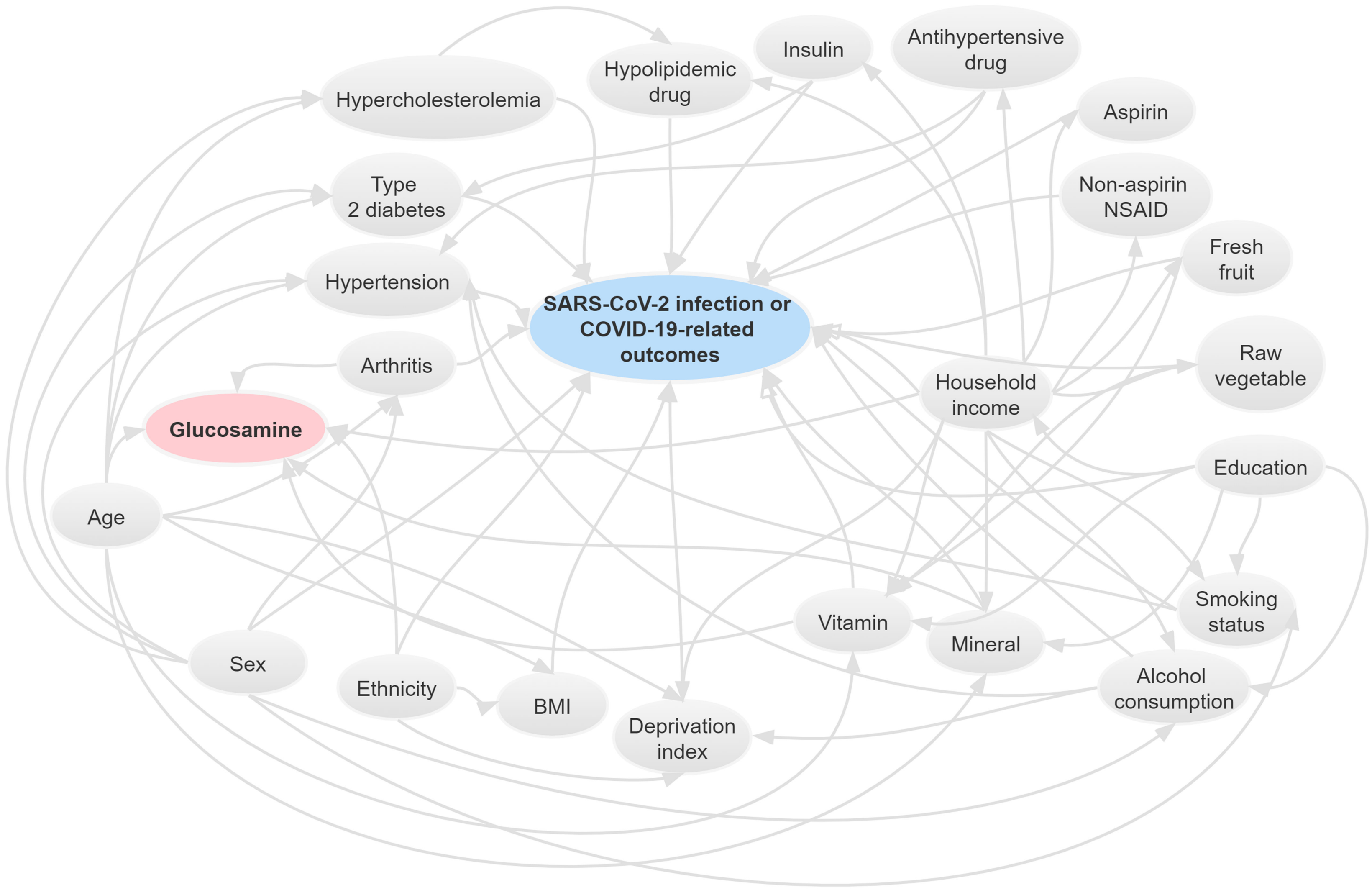
